# Systematic Review and Meta-analysis of the Effects of Transcranial Electric Stimulation on Sleep in Healthy Adults

**DOI:** 10.1101/2025.03.30.25324699

**Authors:** Hiroki Takeuchi, Yuki Motomura, Ayako Imamura, Mitsuaki Takemi, Koichi Hosomi, Akifumi Kishi

## Abstract

**Background:** Transcranial electrical stimulation (tES) has gained attention because of its potential to modulate human sleep physiology. However, its efficacy in healthy populations remains unclear.

**Objectives:** This systematic review and meta-analysis aimed to evaluate the effects of tES on sleep in healthy adults.

**Methods:** A comprehensive literature search was conducted across five electronic databases from January 5 to 10, 2023, to retrieve articles administering tES and evaluating its effects on sleep in healthy adults. The Cochrane Collaboration’s tool was used to assess the risk of bias. The meta-analysis was performed using random-effects models and robust variance estimation, with subgroup analyses and meta-regression to examine conditional and moderation effects.

**Results:** Among the 1,182 identified articles, 39 were included in the qualitative synthesis, and 14 (*N* = 297) met the criteria for meta-analysis. The overall effect of tES on sleep was negligible (estimate: 0.00, 95% confidence interval [CI]: –0.18 to 0.19), with substantial heterogeneity. Subgroup meta-analysis revealed that pre-sleep tES prolonged sleep latency (estimate: –0.30, 95% CI: –0.49 to –0.11), whereas transcranial alternating current stimulation (tACS) increased total sleep time (estimate: 0.14, 95% CI: 0.01 to 0.27). Meta-regression analysis showed that tES efficacy was attenuated with increasing age (*β*: –0.01, 95% CI: –0.02 to 0.00).

**Conclusions:** Although tES does not appear to have a substantial overall effect on sleep, tACS may improve total sleep time, and pre-sleep offline tES may prolong sleep latency. Further well-designed studies are warranted to confirm these findings and minimize potential biases.

## Introduction

Sleep plays a pivotal role in preventing the onset of various health issues, including cardiometabolic diseases [1–4], psychiatric disorders [5,6], and dementia [7,8]. In addition, sleep issues increase the risk of occupational problems, such as work injuries [9,10] and absenteeism [9], leading to substantial economic losses [11,12]. Although habitual sleep patterns vary across countries [13], insufficient sleep and sleep disturbances remain major concerns. A recent meta-analysis estimated that approximately one in four individuals failed to meet age-specific recommended sleep duration (7–9 hours for adults, 6–8 hours for the elderly) [14]. This finding underscores the difficulty of maintaining sufficient sleep in daily life and highlights the potential demand for supportive technologies aimed at improving sleep, even among healthy individuals.

Reflecting such circumstances, products and services, referred to as “sleep technology” (sleep tech), have become increasingly popular in recent years. Their global market is expanding rapidly and is projected to reach USD 95.8 billion by 2032 [15]. Alongside these advancements, products claiming to improve sleep quality through transcranial electrical stimulation (tES) have been developed recently and are available at affordable prices. These claims are often based on the findings from previous non- invasive brain stimulation (NIBS) studies, including transcranial direct current stimulation (tDCS) and transcranial alternating current stimulation (tACS) [16–18]. However, NIBS efficacy in improving sleep quality among healthy individuals remains debatable even in controlled experimental settings. Although some review articles [19–21] have addressed this issue, NIBS effects on sleep have not been quantitatively verified.

Furthermore, tES can be performed with relatively simple equipment compared with other NIBS techniques, such as transcranial magnetic stimulation (TMS) [22] and transcranial focused ultrasound stimulation (tFUS) [23]. This accessibility has led some communities to develop and use their own tES devices for self-improvement [24]. Given the growing interest and accessibility of tES, a fair and comprehensive evaluation of its effects on sleep in healthy individuals is both meaningful and urgent. Such an evaluation will not only be useful for the scientific community but also provide valuable insights for the general public regarding the current status of sleep science and brain stimulation research.

This study aimed to conduct a systematic review and meta-analysis to evaluate the efficacy of tES on sleep in healthy adults and elderly. We also examined the conditional and moderating effects of the experimental and participant characteristics by performing subgroup and meta-regression analyses, to provide a comprehensive overview of the current evidence and several future directions advancing this research topic.

## Methods

This systematic review and meta-analysis followed the Minds Handbook for Clinical Practice Guideline Development 2020 version 3.0 chapter 4 [25]. The review protocol was registered in the International Prospective Register of Systematic Reviews (PROSPERO, CRD42023400091). We completed the PRISMA 2020 checklist [26] to ensure that critical components were included in this paper (Supplementary Material 1).

### Database search

Five journal databases, PubMed, Web of Science, Scopus, PsychINFO, and Ichu-shi (Japanese Medical Manuscript Database), were used to collect articles. Search queries for each database are available in Supplementary Tables 3–7. Briefly, search queries were designed to identify articles with titles and/or abstracts, including terms related to NIBS and sleep. Moreover, we excluded (1) articles written in languages other than Japanese or English, (2) animal studies, and (3) clinical studies. Using these search queries, two authors (YM and AI) independently searched for peer-reviewed articles published up to January 5, 2023.

### Screening procedures

Articles obtained from the databases were screened in two phases. The first screening was performed to remove irrelevant articles based on the relevance of titles and abstracts. Studies involving NIBS other than tES, such as TMS and tFUS, were excluded from the review process. This is because these technologies are not yet accessible to the general public and are beyond the scope of this study.

The second screening was performed by reviewing the full articles on the basis of the following eligibility criteria that contains participant, intervention, comparison, and outcome framework: (1) participants were healthy human adults (age ≥18 years); (2) any form of tES was implemented to modulate subsequent sleep; (3) any type of control condition was employed (e.g., sham-control conditions, no-stimulation conditions, and conditions other than tES); (4) any data that are potentially associated with sleep physiology were measured during and/or after the last stimulation session; (5) the study was an original article published in a scholarly peer-reviewed journal written either in English or Japanese; and (6) the study design was either a randomized controlled trial (RCT) or a non-randomized controlled study (NRS).

Moreover, studies included in the meta-analysis met the following criteria: (7) outcome information was sufficient to calculate the effect size of the intervention, (8) the study design was an RCT, and (9) the study was assessed as having a low or moderate risk of bias and indirectness (see Risk of bias assessment and indirectness assessment). When the screening results differed between YM and AI, other authors (KH, MT, and AK) reviewed the relevant articles to reach a consensus.

### Data extraction

YM and AI independently extracted the following data during the second screening: study design, participant demographics, types of intervention and control conditions, sleep measures, and types of tES. Each experiment in an article was regarded as a single trial. For studies included in qualitative synthesis, YM and AI extracted additional details regarding the interventions and experimental protocols. Moreover, descriptions of adverse events were recorded.

Fundamental statistics (means, standard deviations, and number of participants in each group) on sleep measures after the last tES session from the RCTs were extracted for meta-analysis. Changes in these measures were used as the main outcome if sleep measures were assessed before and after NIBS. For the studies using a crossover design, to avoid possible carryover effects, only the results of the first intervention were analyzed, and subsequent interventions were not included in the analysis. Data from single-condition intervention groups were used in studies using a factorial design. If the fundamental statistics were not available in the manuscript, YM contacted the authors to request additional data. Ultimately, two authors provided the data. The extracted and collected data were entered into a predesigned spreadsheet that automatically calculated the standardized mean difference (SMD) as an effect size. For sleep measures with higher values indicating poor sleep quality (i.e., number of awakenings, wake after sleep onset, duration and ratio of light [N1 and N2] sleep, sleep latency, movement time, fragmentation index, and Pittsburgh Sleep Quality Index score), the sign of SMD was reversed to ensure that greater values consistently represented better sleep.

Sleep measures that reflect similar physiological aspects were often labeled differently across articles. To facilitate subgroup analyses, two authors (YM and AK), experts in human sleep research, classified the collected sleep measures into major categories (i.e., slow-wave [SW] sleep, SW events, spindle events, delta waves, sigma waves, total sleep time, sleep latency, and sleep efficiency).

### Risk-of-bias and indirectness assessment

YM and AI independently conducted the risk-of-bias assessments using the Cochrane Collaboration’s tool [28] for studies with available sleep measurement statistics. Low, high, or unclear risk of bias was determined for each of the following six domains: (1) selection bias, determined by random allocation sequence, baseline imbalance, and allocation concealment; (2) performance bias, determined by blinding of participants and personnel; (3) detection bias, determined by blinding of outcome assessments; (4) attrition bias, determined by the incompleteness of outcome data and intention-to-treat analysis; (5) reporting bias, determined by selective reporting; and (6) other possible sources of bias, such as conflict of interest, a sample size determination method, and incorrect statistical methods. Disagreements were resolved through discussions with five authors (YM, AI, MT, HK, and AK). Studies judged to have a high risk of bias in more than two domains were excluded from meta-analysis.

Additionally, YM and AI independently assessed the indirectness of the evidence for each article in terms of population, intervention, comparison, and outcome. Supplementary Material 2 shows the detailed indirectness assessment criteria and their results. As with the risk of bias assessment, studies judged to have a high overall indirectness were excluded from the meta-analysis.

### Data analysis

For articles that passed the second screening, the study and sample characteristics were tabulated and visualized, including publication year, participants’ age, types of tES, stimulation timing (during wakefulness or sleep, i.e., offline or online), and target brain region. Supplementary Table 8 lists additional characteristics, such as the number of stimulations per day, total stimulation time, stimulation intensity, and stimulation frequency.

For articles that passed the risk of bias and indirectness assessments, the data were compiled into a single dataset for a standard pairwise meta-analysis using a random-effects model. Robust variance estimation was applied to address the nested structure of multiple effect sizes from single trials [29]. Subgroup meta-analyses were performed by stratifying the dataset according to the type of tES, timing of stimulation, stimulation region, age group (adults or elderly), sleep variables, and study design (parallel or crossover). Additional stratified analyses were performed beyond the pre-registered protocol when the results were likely to be influenced by other experimental characteristics. Heterogeneity across the effect sizes was assessed using Cochran’s *I^2^* and *Q* statistics. The imprecision of evidence was determined based on optimal information size and confidence intervals [30]. The possibility of publication bias was examined using Begg’s and Egger’s tests and visualized using a funnel plot. Certainty of evidence was evaluated using the grading of recommendations, assessment, development, and evaluation (GRADE) methodology [31], rating relevant five factors (risk of bias, indirectness, inconsistency, imprecision, and publication bias) as high (–2), moderate (–1), or low (0). The rating procedures are described in Supplementary Material 2.

Furthermore, random-effect meta-regression analyses were performed to examine the relationship between tES efficacy and factors, such as stimulation frequency (particularly for tACS), stimulation intensity, size of the stimulation electrode, number of stimulations per day, and total stimulation time. Participants’ age data were also used for the meta-regression analysis to examine linear relationships with the obtained effect size.

To confirm the robustness of the findings obtained through the random-effects meta-analysis, similar analyses were performed using a fixed-effects model. All analyses were performed using the R statistical software (version 4.0.1). Pairwise meta-analyses, meta-regressions, and visualizations were performed using the metafor package.

## Results

### Search results

Fig. 1 shows the literature review process. We identified 1,811 articles from the five databases and one additional article from the reference sections of the retrieved studies. After excluding 1,050 articles owing to irrelevant research aims based on their titles and abstracts, 132 articles proceeded to a full-text eligibility assessment. Among these, 93 articles were excluded from the second screening for the following reasons: studies that included patient populations (*n* = 50), studies that did not report the results of sleep measurements (*n* = 25), articles that were not original or peer-reviewed (*n* = 14), and studies that did not use tES (*n* = 2). One duplicate study (*n* = 1) and one secondary analysis that did not focus on effects on sleep (*n* = 1) were excluded. Full texts from the remaining 39 articles [32–70] were included in the qualitative synthesis. Finally, 14 RCT articles [57–70] were included in the meta-analysis. The reasons for exclusion were as follows: the studies were NRSs (*n* = 13), the reported statistical values were insufficient to calculate effect sizes (*n* = 7), and the studies had a high risk of bias (*n* = 4). Additionally, one study was excluded because the reported results may have been influenced by experimental manipulations unrelated to tES (high indirectness).

**Figure 1.**
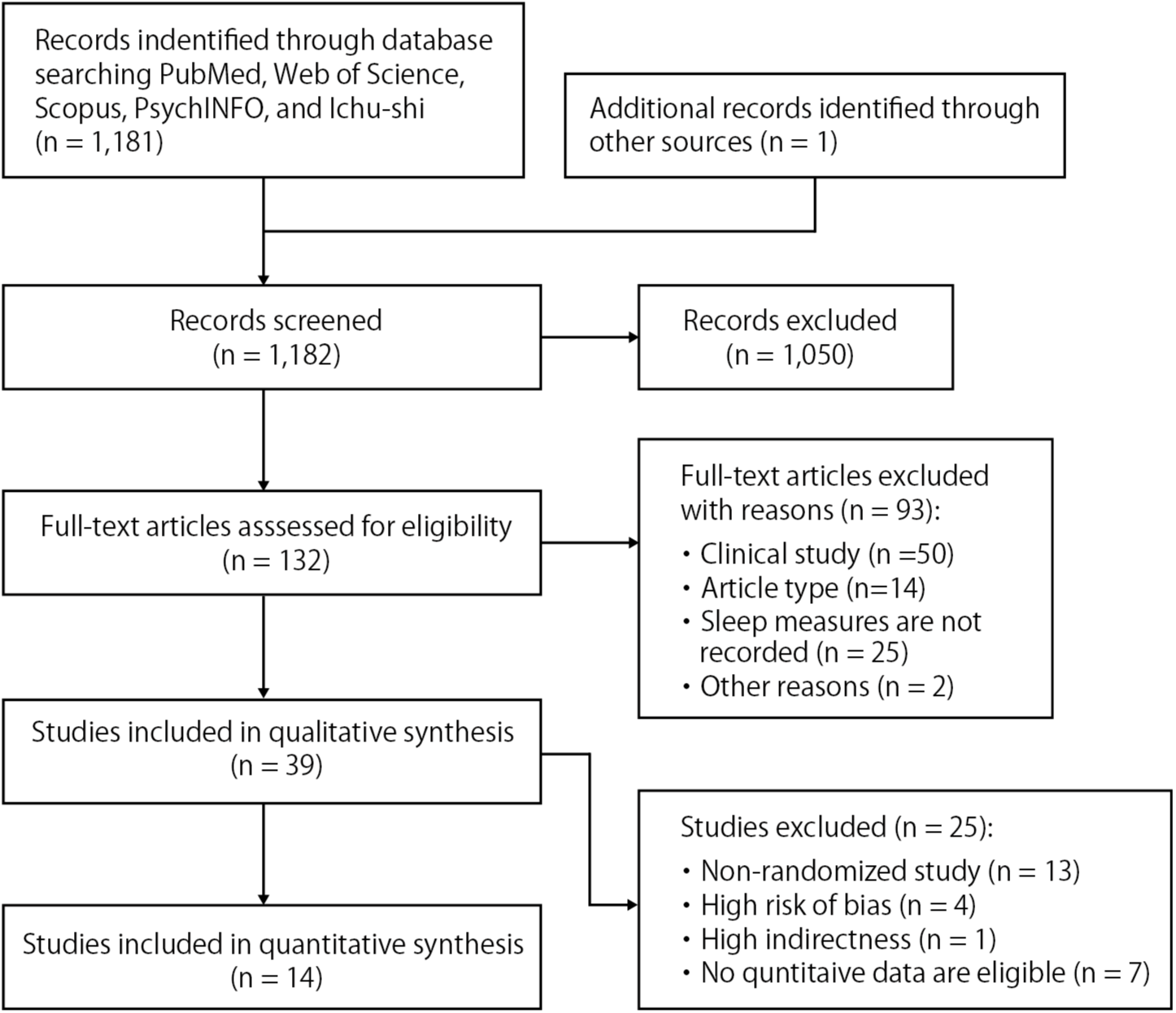
Flowchart of the article screening process.

### Study and sample characteristics

Fig. 2 provides an overview of the results of the qualitative synthesis. The publication years of the screened articles ranged from 2004 to 2022, with more than half (25/39) of them published after 2017 (Fig. 2A). The tES types in included studies were as follows (Fig. 2B): tACS in 12 experiments (offline:online = 3:9); static tDCS in 17 experiments (offline:online = 13:4); slow-oscillatory tDCS (so-tDCS) and theta-wave tDCS (theta-tDCS) during sleep in 13 and 3 experiments, respectively; and cranial electrotherapy stimulation (CES) in 3 experiments (offline:online = 2:1). The other two experiments included a study that used unique stimulation methods not classified under the general tES category and one study in which the type of stimulation was indeterminate. Seven studies were conducted on tES in an elderly population (≥65 years, Fig. 2C). Regarding the targeted brain regions, most studies focused on the frontal regions (e.g., F3 and F4 in the 10-20 system, Figs. 2D and 2E). More detailed information, including stimulation intensity, frequency, and duration, is provided in Supplementary Table 8.

**Figure 2.**
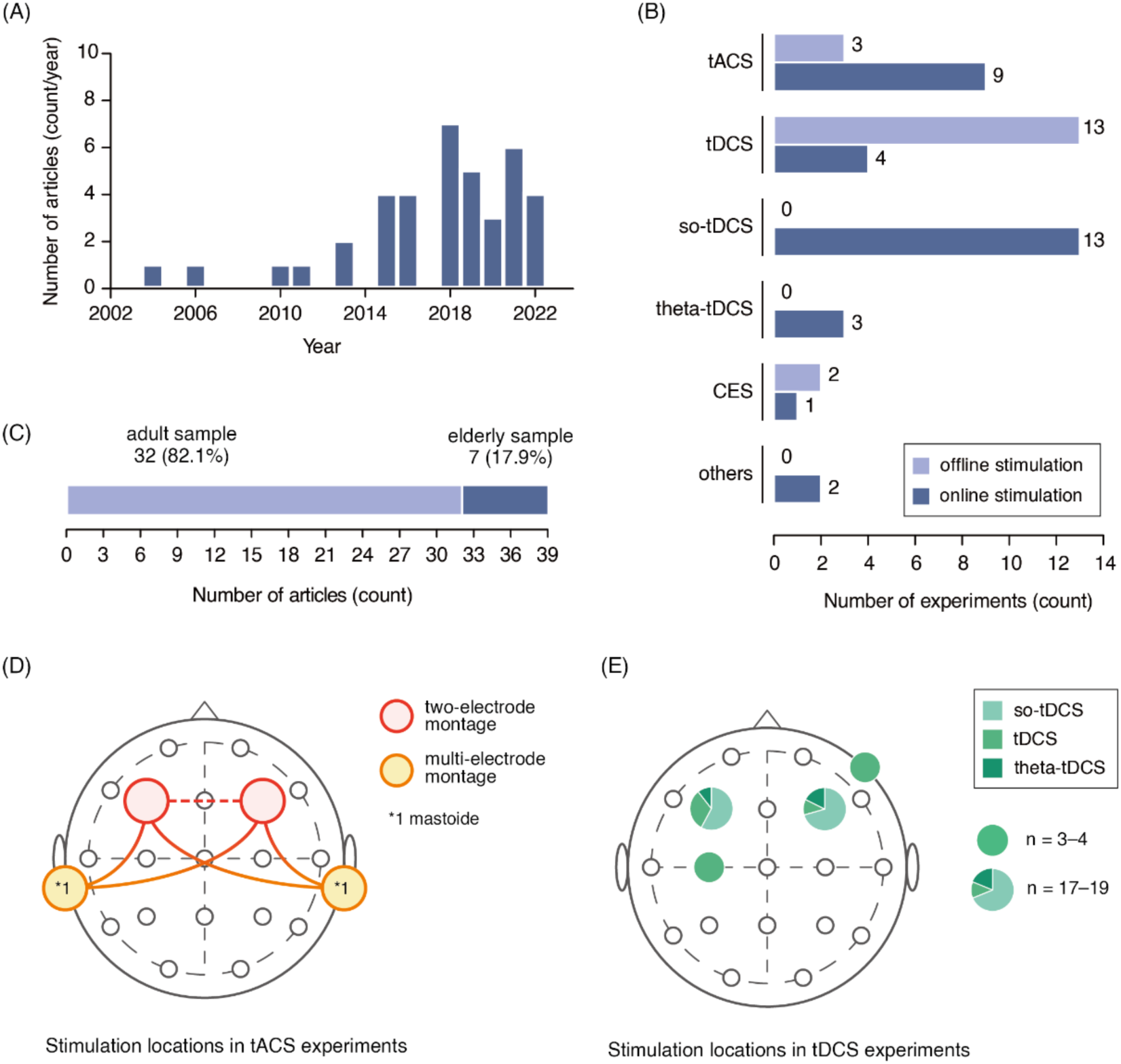
Overview of the study characteristics. (A) Publication years of the included articles. (B) Number of studies stratified based on stimulation type and timing of stimulation. (C) Participant age groups. (D) Electrode placement in transcranial alternating current stimulation (tACS) trials. (E) Electrode placement in transcranial direct current stimulation (tDCS) trials. Note that (D) and (E) show only electrode placements employed in three or more trials. Abbreviations: so-tDCS, slow oscillatory tDCS; theta-tDCS, theta wave tDCS; CES, cranial electrotherapy stimulation.

### Adverse events and side effects

Twelve articles investigated adverse events or side effects related to electrical stimulations (Supplementary Table 8), with five reporting adverse events. Specifically, a tingling sensation was reported by six participants [55], stinging sensation by one [55], nausea by one participant [69], unpleasant physical sensation by one [38], and superficial skin lesion by one participant [60]. One study did not provide the specific incidence data [57].

Only one study [60] quantitatively assessed the frequency of adverse events in 19 participants. According to their report, participants felt prickling (*n* = 17), itching (*n* = 9), and burning (*n* = 4) sensations on the skin under stimulation electrodes with frequencies comparable to those in the control condition. However, the rate of reported headaches in the tES condition was approximately twice than that in the control condition (tES condition, *n* = 5; control condition, *n* = 2).

### Risk-of-bias and indirectness assessment

Following qualitative synthesis, articles were further screened based on the availability of quantitative sleep measurements and the inclusion of control conditions. The risk of bias and indirectness of evidence were assessed for the remaining 19 articles. The overall risk of bias was high in 4 articles and moderate in 15 articles (Fig. 3). In the assessment of the indirectness of evidence, only one study was rated as having high overall indirectness (Supplementary Material 2). Fourteen articles that passed both the risk of bias and indirect assessments were included in the following meta-analyses.

**Figure 3.**
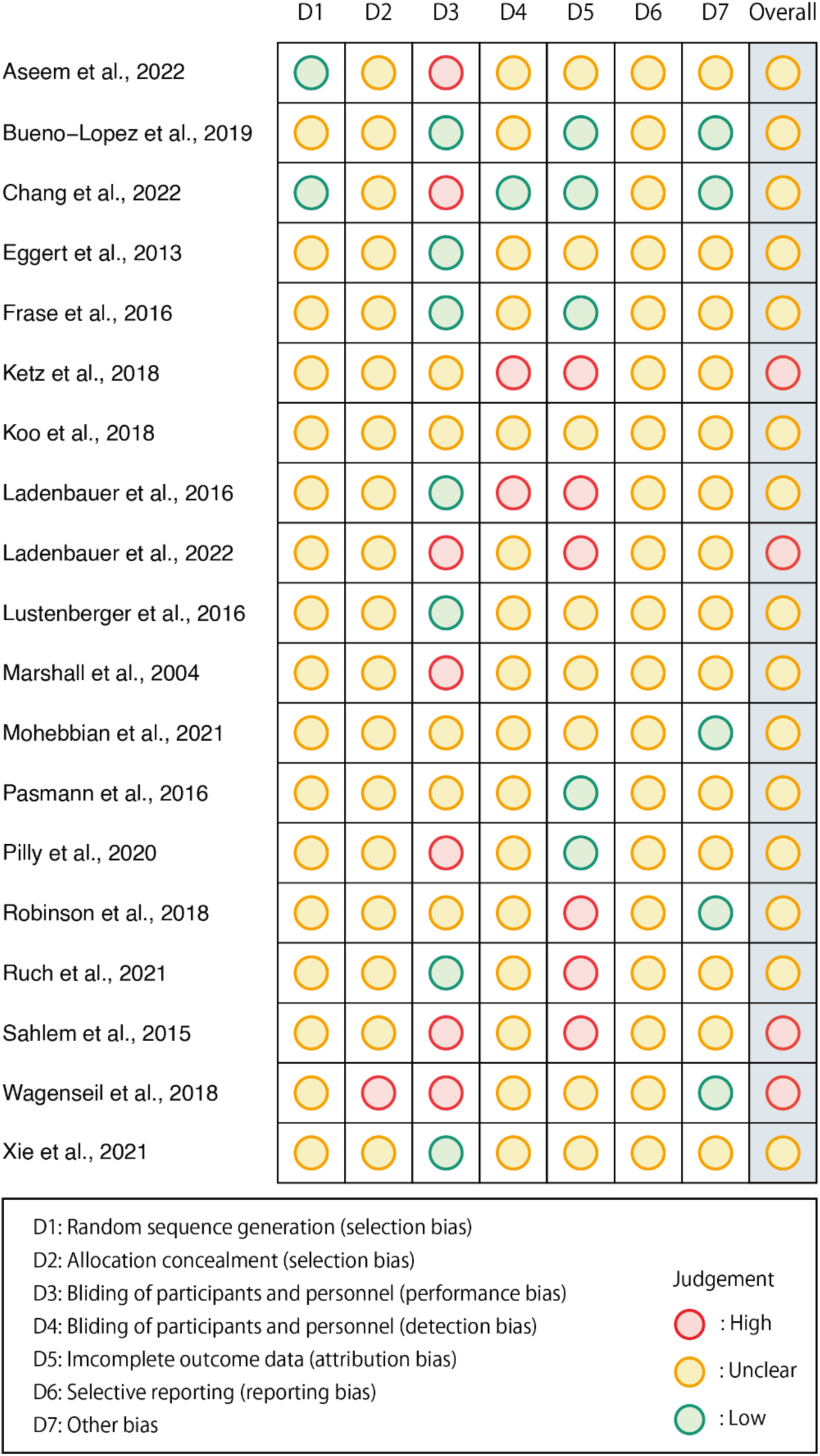
Summary of the risk-of-bias assessment. Green, yellow, and red circles indicate low, unclear, and high risk of bias, respectively, in each assessment category.

### General effects of tES on sleep

To evaluate the efficacy of tES on sleep, a random-effects meta-analysis (total sample size = 297) was performed for the SMDs of the sleep variables. The result indicated that the general effect of tES on sleep was close to null (estimate = 0.00, 95% confidence interval [CI] = [–0.18, 0.19], *p* = 0.96; Fig. 4A) with considerable heterogeneity (*I^2^*= 79.2%, *Q* = 866.2, *p* for *Q* < 0.01). Egger’s and Begg’s tests showed a significant slope (*z* = 5.76, *p* < 0.01) and rank correlation (*τ* = 0.12, *p* = 0.01) between the effect size and sampling variances, suggesting the existence of publication biases. The funnel plot supported these results (Fig. 5). To ensure the robustness of our findings, we performed a similar analysis using a fixed-effects model. Consequently, the fixed effect meta-analysis replicated the negligible general effect (estimate = –0.02, 95% CI = [–0.17, 0.12], *p* = 0.72; Supplementary Tables 9 and 10).

**Figure 4.**
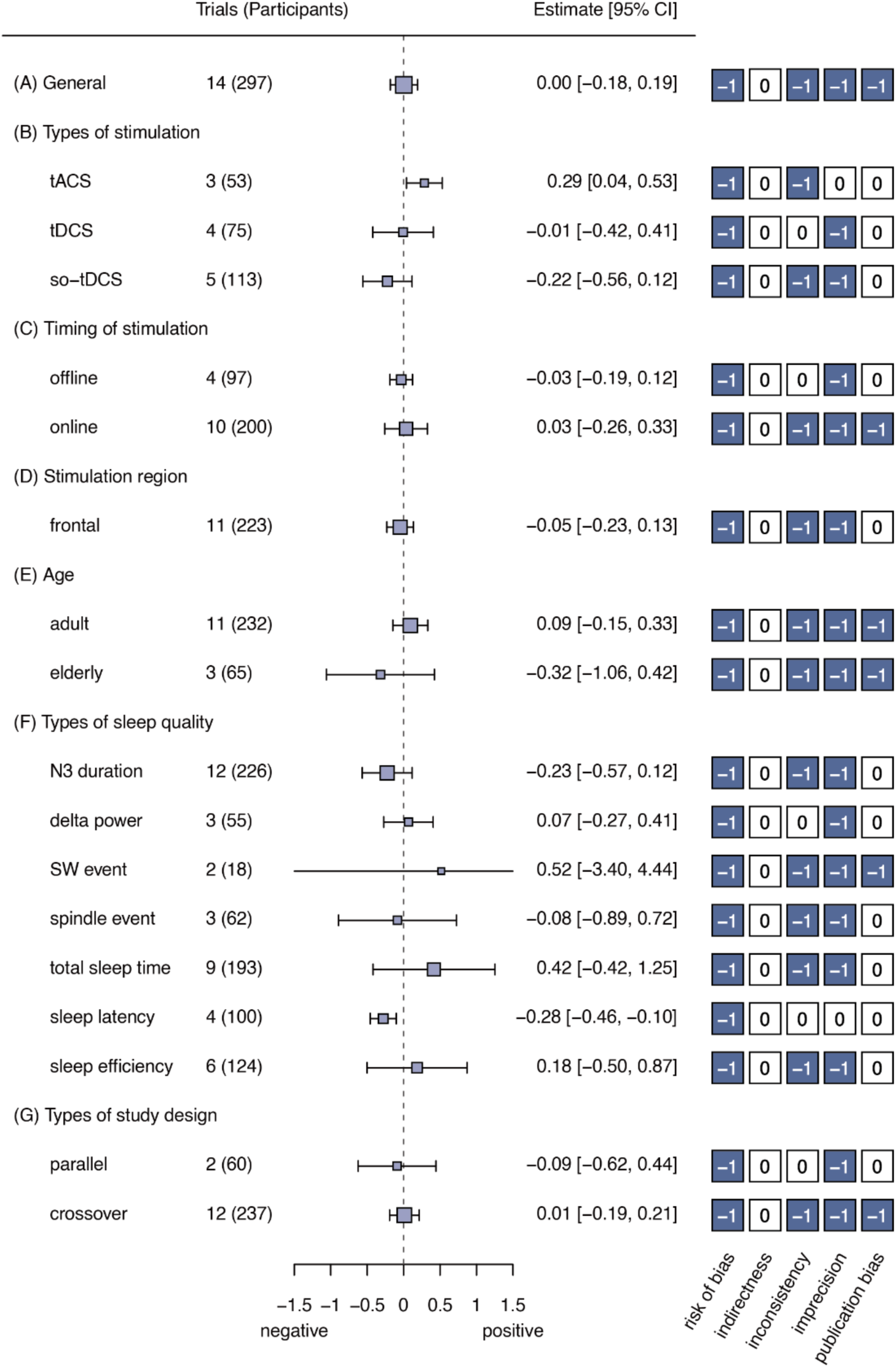
Forest plots for overall and subgroup meta-analyses. The numbers -1 and 0 in the boxes next to the forest plots represent moderate and low risks, respectively, for factors that reduce the certainty of evidence. Abbreviations: tACS, transcranial alternating current stimulation; tDCS, transcranial direct current stimulation; so-tDCS, slow oscillatory transcranial direct current stimulation; SW, slow wave.

**Figure 5.**
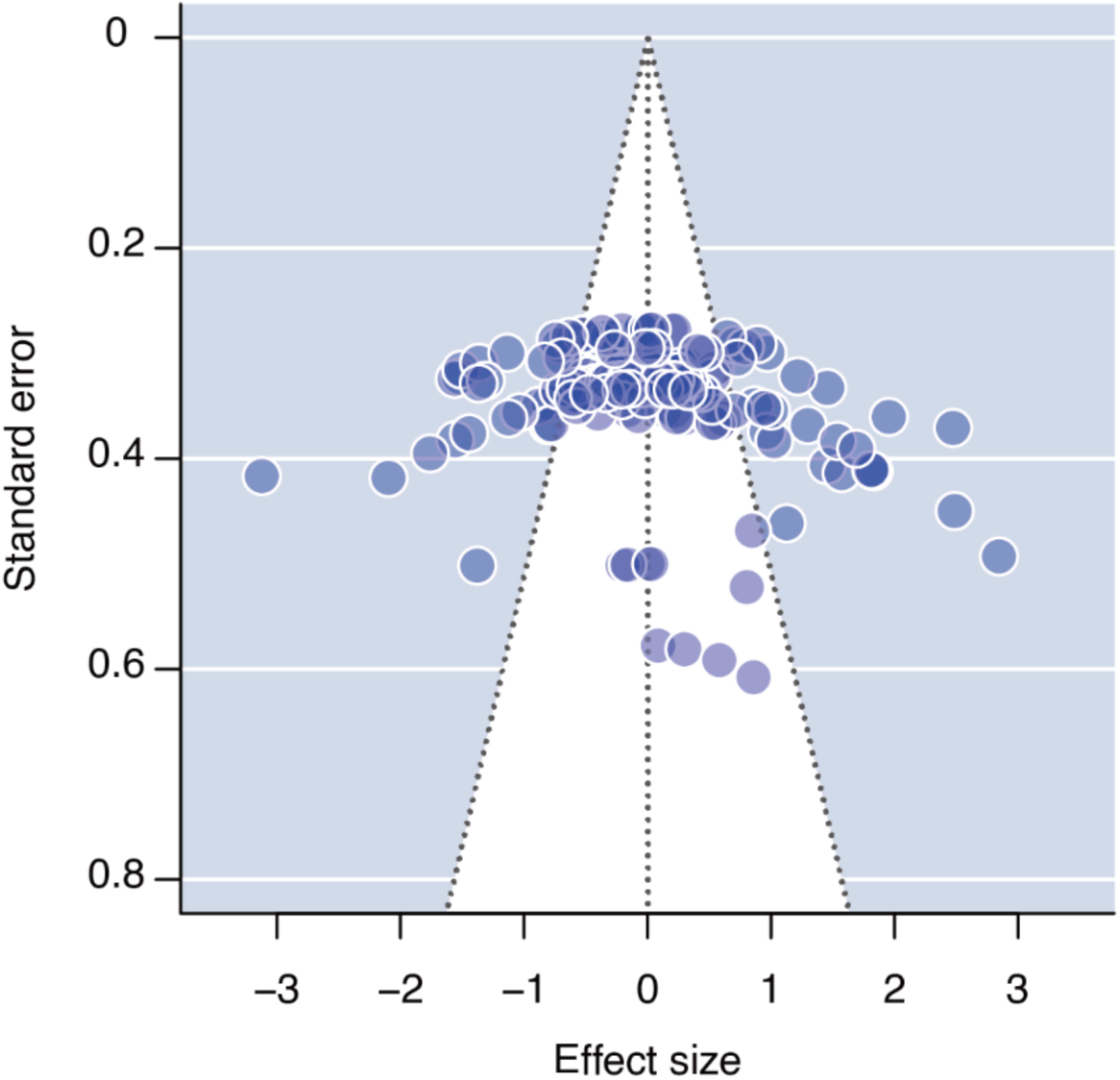
Funnel plot for the overall meta-analysis. Blue points indicate the collected effect sizes.

### Subgroup meta-analyses

Subgroup meta-analyses were performed to examine the conditional effects of the experimental and participant characteristics. When stratified by type of stimulation (tACS, tDCS, and so-tDCS; Fig. 4B), tACS demonstrated a small and significant positive effect on subsequent sleep (estimate = 0.29, 95% CI = [0.04, 0.53], *p* = 0.04); nevertheless, the heterogeneity remained substantial (*I^2^* = 60.5%, *Q* = 77.8, *p* for *Q* < 0.01). In contrast, tDCS showed no significant effect on sleep (estimate = –0.01, 95% CI = [–0.42, 0.41], *p* = 0.92), without heterogeneity (*I^2^* = 12.1%, *Q* = 86.5, *p* for *Q* = 0.19). so-tDCS lacked a significant effect (estimate = –0.22, 95% CI = [–0.56, 0.12], *p* = 0.14), with considerable heterogeneity (*I^2^*= 86.0%, *Q* = 492.3, *p* for *Q* < 0.01). Owing to the limited number of trials, analyses were not performed for the CES or other stimulation types (each with only one trial). When stratified by sleep variable (Fig. 4F), only sleep latency was significantly affected (prolonged) by administering tES (estimate = –0.28, 95% CI = [–0.46, –0.10], p = 0.02, *I^2^* = 24.6%, *Q* = 9.3, *p* for *Q* = 0.23). Notably, the sign of SMD for sleep latency was reversed so that the negative values consistently represented poor sleep quality in this study. Stratification by other experimental characteristics did not reveal any significant effects of tES on sleep (Figs. 4C, D, E, and G).

### Additional analyses

Given the positive effect of tACS on sleep in general, with considerable heterogeneity of the effect, as indicated earlier, further analysis was performed to clarify which specific aspects of sleep were modified by tACS. To resolve this question, the efficacy of tACS was further examined by stratifying sleep variables (Fig. 6A, Supplementary Tables 11 and 12). Owing to the number of tACS trials, the analysis focused on N3 duration, total sleep time, and sleep efficiency. The results indicated that tACS significantly increased total sleep time (estimate = 0.14, 95% CI = [0.01, 0.27], p = 0.05, *I^2^* = 0.0%, *Q* = 0.92, *p* for *Q* = 0.63). However, no significant effects were found for N3 duration (estimate = –0.15, 95% CI = [–0.70, 0.40], p = 0.37, *I^2^*= 0.0%, *Q* = 0.92, *p* for *Q* = 0.63) or sleep efficiency (estimate = 0.66, 95% CI = [–2.33, 3.65], p = 0.22, *I^2^* = 62.7%, *Q* = 8.3, *p* for *Q* = 0.04).

**Figure 6.**
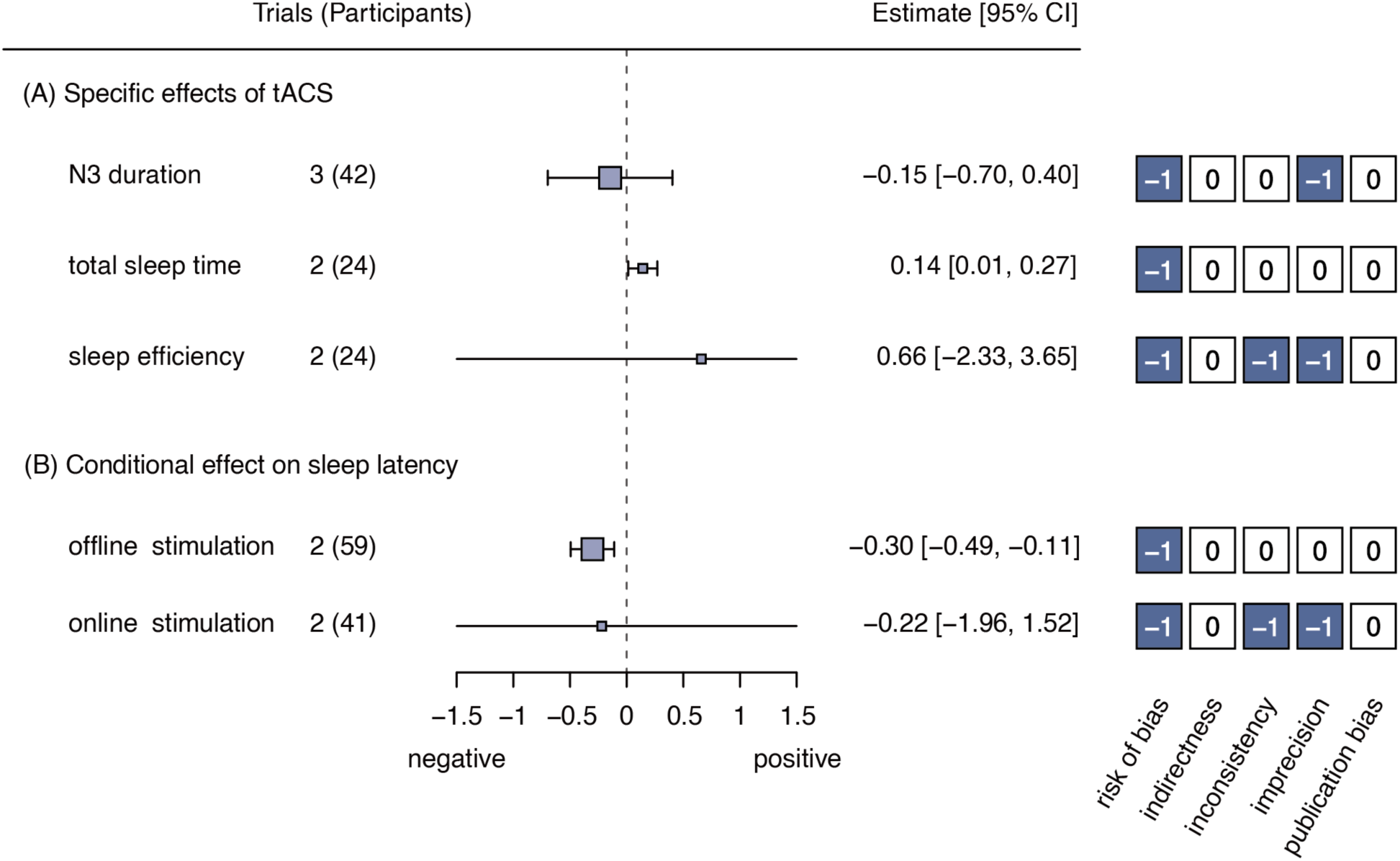
Forest plots for additional analyses. The numbers -1 and 0 in the boxes next to the forest plots represent moderate and low risks, respectively, for factors that reduce the certainty of evidence. Abbreviation: tACS, transcranial alternating current stimulation.

In online stimulation protocols, stimulation is provided after sleep onset. Thus, the aforementioned negative effects on sleep latency may be conditioned by the stimulation timing. To investigate this possibility, the data were stratified according to whether stimulations were presented during wakefulness or sleep (online or offline), followed by pairwise meta-analyses for each condition (Fig. 6B, Supplementary Tables 11 and 12). This subset included eight trials (tACS:tDCS:so-tDCS:CES = 2:4:1:1). Consequently, the results indicated that sleep latency was significantly prolonged with offline protocols (estimate = –0.30, 95% CI = [–0.49, –0.11], p = 0.03, *I^2^* = 0.0%, *Q* = 3.6, *p* for *Q* = 0.46). In contrast, online stimulation did not significantly affect sleep latency (estimate = –0.22, 95% CI = [–1.96, 1.52], p = 0.36, *I^2^* = 75.3%, *Q* = 34.8, *p* for *Q* < 0.01).

### Meta-regression analysis

Meta-regression analyses were performed to examine the relationships between the effect size and experimental characteristics that can be expressed as continuous values, such as stimulation frequency, stimulation intensity, size of the stimulation electrode, number of stimulations per day, and total stimulation time during the study period. No significant relationships were found between these experimental characteristics and the effect sizes (Fig. 7 and Supplementary Tables 13 and 14). However, a meta-regression analysis found a significant negative relationship between age and the effect size (*β* = –0.01, 95% CI = [–0.02, 0.00], *p* = 0.03), suggesting that the efficacy of tES was attenuated with increasing age.

**Figure 7.**
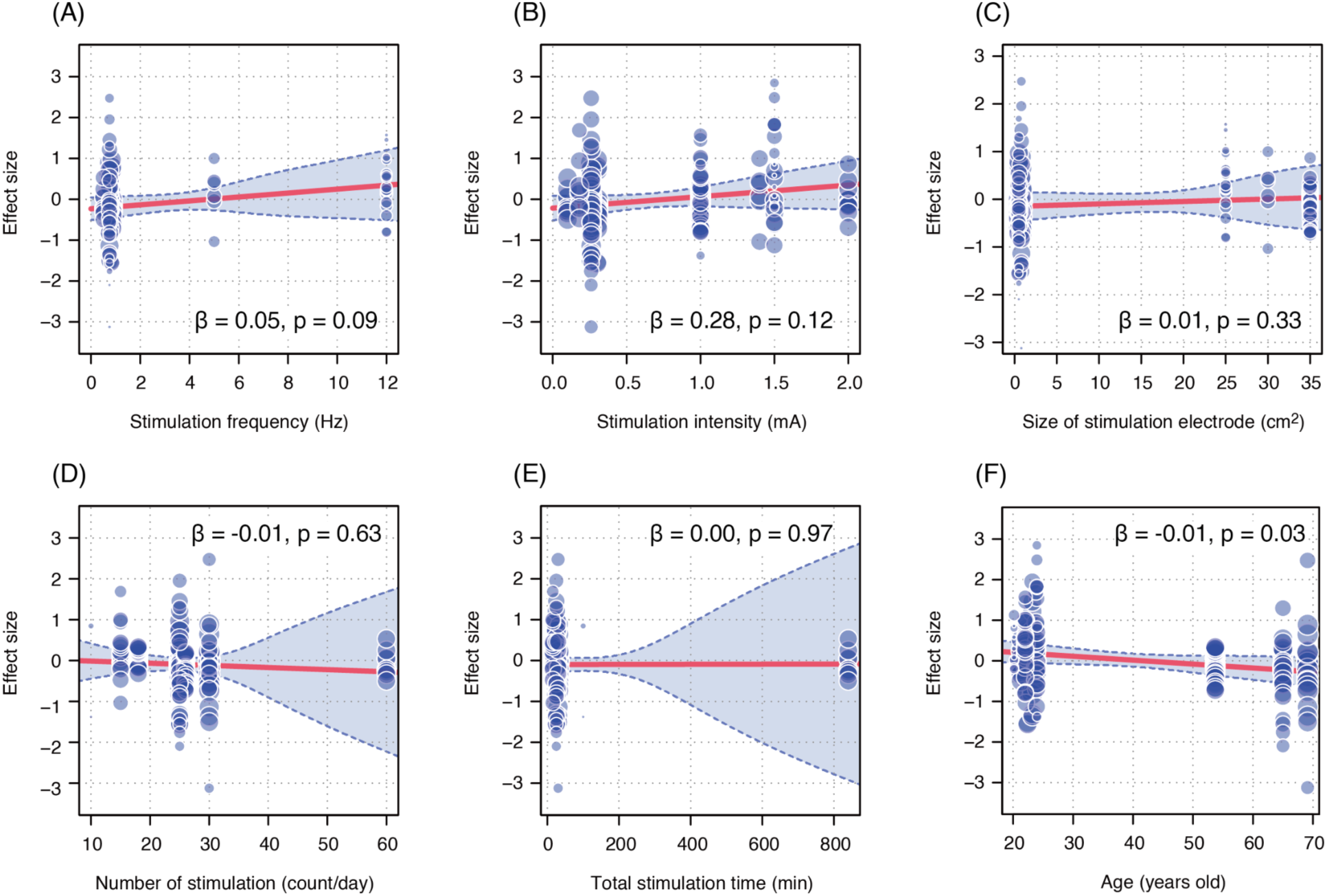
Meta-regression analyses. The scatterplots show the relationship between the collected effect size (blue points) and various experimental parameters: (A) stimulation frequency, (B) stimulation intensity, (C) electrode size, (D) number of stimulations per day, (E) total stimulation time over the trial period, and (F) participants’ mean age per trial. Red lines and shaded blue areas indicate regression lines and 95 % confidence intervals, respectively. β and p values in each plot indicate the regression coefficients and statistical significance, respectively.

## Discussion

To our knowledge, this is the first study to evaluate tES effects on sleep in healthy populations using both a systematic review and meta-analysis. Although a previous systematic review [71] explored similar issues, it focused on patients with neurological and psychiatric disorders and included uncontrolled and quasi-experimental studies with a high risk of bias. Additionally, evidence concerning healthy populations has been limited to narrative reviews [19,20] and qualitative synthesis [21].

We used a two-phase screening process to exclude articles with a high risk of bias and indirectness before conducting quantitative evidence synthesis. Eventually, 39 articles were identified for qualitative synthesis, and 14 articles were included in the meta-analysis. The overall meta-analysis revealed that tES effect on sleep was close to null with considerable heterogeneity in the reported effect sizes. Moreover, Egger’s and Begg’s tests suggested a potential publication bias. These findings indicate that tES effects on sleep remain unclear. However, considering the recent growth in the sleep tech market, it is important to clarify the current status of evidence regarding the effects of tES on sleep in healthy populations. Our findings provide critical insights for the general public regarding consumer-grade NIBS (especially tES) device usage to improve sleep. Importantly, this study focused on previous tES studies conducted in experimental settings and does not dismiss the potential utility of specific commercial NIBS devices in real-world settings.

The negligible effect observed in the general meta-analysis may be attributed to methodological diversity. This analysis aggregated all the collected effect sizes without stratification by experimental characteristics, including the type and timing of the stimulation (online or offline). Cortical activity rhythms dramatically differ between wakefulness and sleep. In electroencephalograms, alpha and beta band activities (8–12 Hz and 16–30 Hz) dominate during wakefulness, whereas delta band activity is prominent during deep sleep (SW sleep). These differences may obscure the effects of stimulation. Furthermore, three of the 14 articles included in the meta-analysis [62,64,69] recorded polysomnography during short-term naps (< 3 h), potentially limiting the assessment of stimulation effects on deeper sleep stages. Future studies should include more nocturnal sleep experiments to better understand tES effects on the overall sleep architecture.

### Findings from subgroup meta-analyses and meta-regression analyses

Although the overall meta-analysis revealed the null effect of tES on sleep, subsequent analyses provided several noteworthy findings that could help advance future brain stimulation research. First, a subgroup meta-analysis suggested that tACS positively modulated overall sleep, primarily by increasing total sleep time. tACS delivers alternating electrical currents to specific brain regions, entraining neuronal firing rates, and modulating cortical rhythms [16,17]. In our analysis, all tACS trials used alternating currents with frequencies at most below the alpha band and targeted the frontal regions (F3, F4, F7, and F8). Given that slow-wave brain activity originates in the anterior areas during sleep onset [72,73], tACS of the frontal regions may promote sleep consistency by slowing the cortical activity rhythm. However, considering the small effect size (estimate = 0.14; Fig. 6A), it remains unclear whether regular use of the current tACS protocols yields an improvement in total sleep time that benefits human health.

Additional analyses showed that pre-sleep (offline) stimulation prolonged sleep latency. Notably, four of the five effect sizes in this analysis came from tDCS trials that targeted the frontal cortex. One study not included in this meta-analysis compared the effects of tDCS and caffeine on psychological and neurobehavioral performance in sleep-deprived individuals [74] and found that 30-min anodal tDCS to the frontal region (F3, 2 mA) prevented an impairment of vigilance performance and improved subjective symptoms (e.g., reduced fatigue and drowsiness) compared with 200 mg caffeine intake. Such excitatory effects of tDCS may interfere with sleep initiation. However, inconsistent findings have been reported [75], and the effects of tES on neurobehavioral functions, particularly wakefulness, remain inconclusive. Further studies are required to comprehensively understand the effects of tES on sleep– wake regulatory systems in humans.

Although subgroup meta-analyses found negligible effects of tES in both adult and elderly populations, a meta-regression analysis showed a negative relationship between age and effect size, suggesting that neuronal aging may reduce the efficacy of tES. Aging is associated with functional and structural changes in the brain, including altered resting-state cortical rhythms [76,77], regional shrinkages in the cortex [78], and ventricular enlargement [79]. These changes may influence cortical electric field distribution, diminishing tES efficacy. Future studies should consider age-related neuronal changes when optimizing the stimulation parameters. Meanwhile, total sleep time generally decreases with age, primarily because of reduced SW sleep [80]. Frequent awakening and reduced sleep efficiency are common concerns among the elderly [80]. Moreover, a previous meta-analysis [7] suggested that insufficient and poor sleep quality increased the risk of neurodegenerative diseases. Thus, older individuals represent a population in need of improved sleep. The development of novel NIBS methodologies to improve sleep in this demographic can significantly contribute to promoting a healthy life expectancy.

### Limitations

This study has some limitations. First, the number of articles included in the meta-analysis (14 articles) was considerably smaller than that in the systematic review (39 articles). Only RCT studies with moderate-to-low risk of bias and indirectness were included in the meta-analysis. Notably, 30% of the articles included in the systematic review were NRS (13/39). Even among the randomized controlled designs, some were excluded because of the potential risks of attribution or detection bias. This highlights the need for more well-designed studies with pre-registered trials to minimize bias.

Second, we could not examine the middle- to long-term effects of tES on sleep because most studies (37/39) recorded sleep only immediately after stimulation. Only a few studies [49,58] have used wearable sleep-monitoring technologies, including actigraphy, to evaluate the stimulation effects. Caution is required when extrapolating these findings into daily-life situations.

Third, in qualitative synthesis, fewer than half of the studies examined adverse events following tES administration, limiting our ability to comprehensively evaluate its safety. Although adverse events from tES are considered rare under controlled conditions as typical stimulation intensities are approximately 2 mA [24], systematic evaluations comparing the frequency of adverse events between the stimulation and control conditions are necessary.

Finally, our inclusion criteria excluded TMS and tFUS experiments, focusing solely on tES. Further studies are required to investigate whether other stimulation techniques also have beneficial effects on sleep physiology.

## Conclusions

This systematic review and meta-analysis evaluate tES effects on sleep in healthy adults. Although the general effect was negligible, subgroup analyses indicated that tACS might improve total sleep time, whereas pre-sleep offline tES could prolong sleep latency. Meta-regression analysis further suggested that tES efficacy diminishes with age. These findings highlight the complexity of tES effects on sleep. Further well-designed studies are warranted to refine stimulation protocols, investigate long-term effects, and evaluate potential applications for improving sleep quality in patient populations.

## Data and code availability

The fundamental statistics collected for each study (i.e., mean, standard deviation, sample size) and analysis codes are accessible on the OSF website (https://osf.io/jznce/).

## CRediT authorship contribution statement

**Hiroki Takeuchi:** Validation, Visualization, Writing - original draft, Writing - review & editing.

**Yuki Motomura:** Investigation, Data Curation, Formal analysis, Writing - review & editing.

**Ayako Imamura:** Investigation, Data Curation, Formal analysis, Writing - review & editing.

**Mitsuaki Takemi:** Conceptualization, Writing - review & editing, Project administration, Funding acquisition.

**Koichi Hosomi:** Methodology, Investigation, Writing - review & editing, Supervision.

**Akifumi Kishi:** Conceptualization, Methodology, Investigation, Writing - review & editing, Supervision, Project administration.

## Acknowledgements

This work was conducted as a part of NeuroTech Guidebook development in JST Moonshot R&D to MT (Grant Number JPMJMS2012). We thank the members of the Evidence Evaluation Committee, a specially organized group for the Moonshot R&D project, for their comments on the manuscript.

