## Supplementary Material 2 for "Systematic Review and Meta-analysis of the Effects of Transcranial Electric Stimulation on Sleep in Healthy Adults"

### Supplementary Material 2: Additional information of quality assessment

#### 1. Criteria and results of indirectness assessment

Five authors (YM, AI, MT, KH, and AK) established criteria for indirectness assessment in terms of population, intervention, comparison, and outcome (Supplementary Table 1). Two authors (YM and AI) independently assessed the indirectness of evidence for the screened 19 RCT studies (Supplementary Table 2). One study was rated as high overall indirectness and excluded meta-analysis.

#### 2. Assessment of certainty of evidence

To provide information related to certainty of evidence, five relevant factors (risk of bias, indirectness, inconsistency, imprecision, and publication bias) were rated as high (-2), moderate (-1), or low (0) for each pair-wise meta-analysis. The assessment followed the grading of recommendations, assessment, development, and evaluation (GRADE) methodology [S1]. Rating procedures for each factor were illustrated in Supplementary Figure 1–5. Optimal information sizes (OISs) were calculated based on Watterslev et al. (2009) [S2].

**Supplementary Table 1: Assessment criteria for indirectness**

| | **Low** | **High** | **Unclear** |
|---|---|---|---|
| Population | Studies targeting healthy adults or older adults | Studies including individuals who have sleep problems (even if they don't have a sleep disorder) | no description |
| Intervention | Studies administering transcranial electrical stimulation (tES) | Studies that cannot be ruled out the effects other than tES | unclear description |
| Comparison | Studies recording (objective and subjective) sleep data | Studies measuring data different from sleep physiology (e.g., fatigue, etc.) | unclear description |
| Outcome | Studies numerically assessing the side effects of tES | | no description |

**Supplementary Table 2: Results of indirectness assessment**

| Article | Population | Intervention | Comparison | Outcome | Overall |
|---|---|---|---|---|---|
| Ruch et al., 2021 | **L** | **L** | **L** | **L** | **L** |
| Pasmann et al., 2016 | **L** | **L** | **L** | **L** | **L** |
| Ladenbauer et al., 2016 | **L** | **L** | **L** | **L** | **L** |
| Ketz et al., 2018 | **L** | **H** | **L** | **L** | **L** |
| Aseem et al., 2022 | **H** | **H** | **H** | **L** | **H** |
| Chang et al., 2022 | **H** | **L** | **L** | **L** | **L** |
| Koo et al., 2018 | **L** | **L** | **L** | **L** | **L** |
| Lustenberger et al., 2016 | **L** | **L** | **L** | **L** | **L** |
| Xie et al., 2021 | **L** | **L** | **L** | **L** | **L** |
| Frase et al., 2016 | **L** | **L** | **L** | **L** | **L** |
| Eggert et al., 2013 | **L** | **L** | **L** | **L** | **L** |
| Pilly et al., 2020 | **L** | **L** | **L** | **L** | **L** |
| Sahlem et al., 2015 | **L** | **L** | **L** | **L** | **L** |
| Bueno-Lopez et al., 2019 | **L** | **L** | **L** | **L** | **L** |
| Robinson et al., 2018 | **L** | **L** | **L** | **L** | **L** |
| Wagenseil et al., 2018 | **L** | **L** | **L** | **L** | **L** |
| Mohebbian et al., 2021 | **L** | **L** | **L** | **L** | **L** |
| Ladenbauer et al., 2022 | **L** | **L** | **L** | **L** | **L** |
| Marshall et al., 2004 | **L** | **L** | **L** | **L** | **L** |

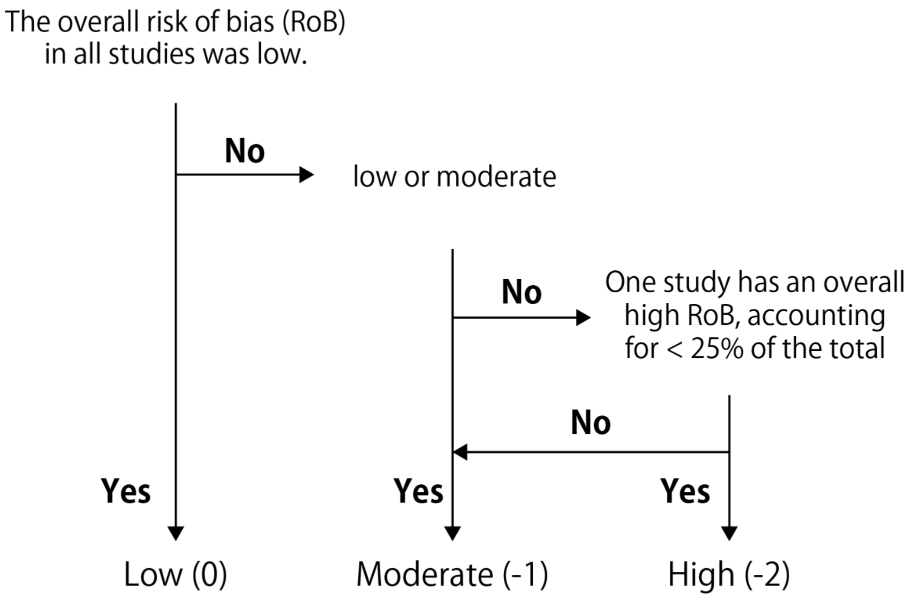


**Supplementary Figure 1.** Rating procedure for risk of bias.

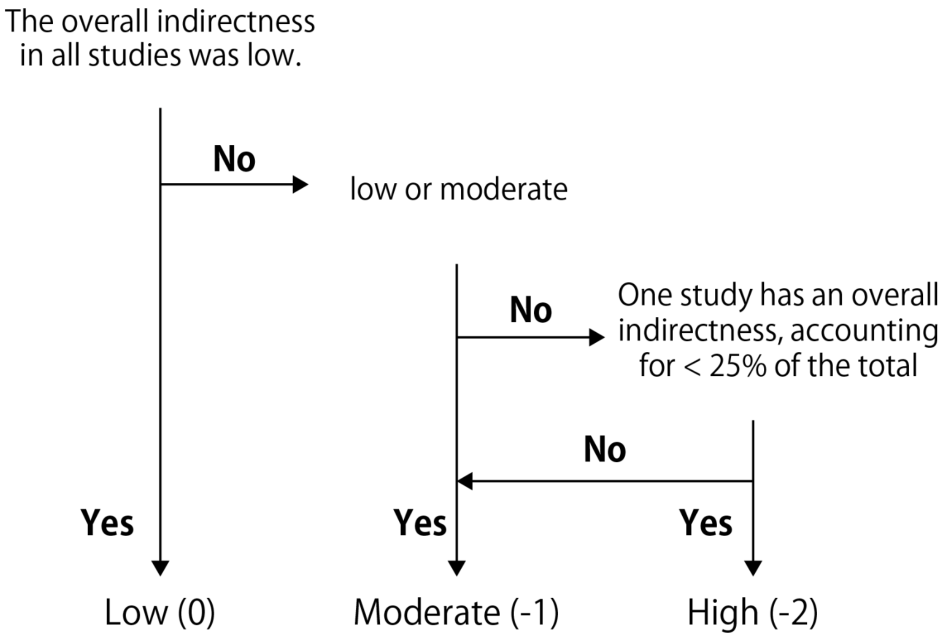


**Supplementary Figure 2.** Rating procedure for indirectness.

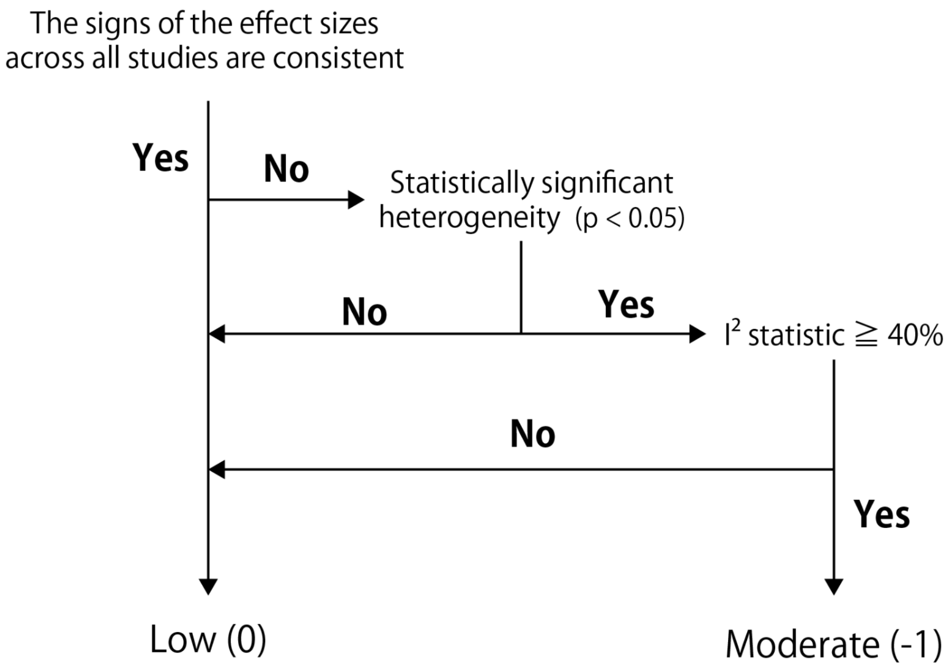


**Supplementary Figure 3.** Rating procedure for inconsistency.

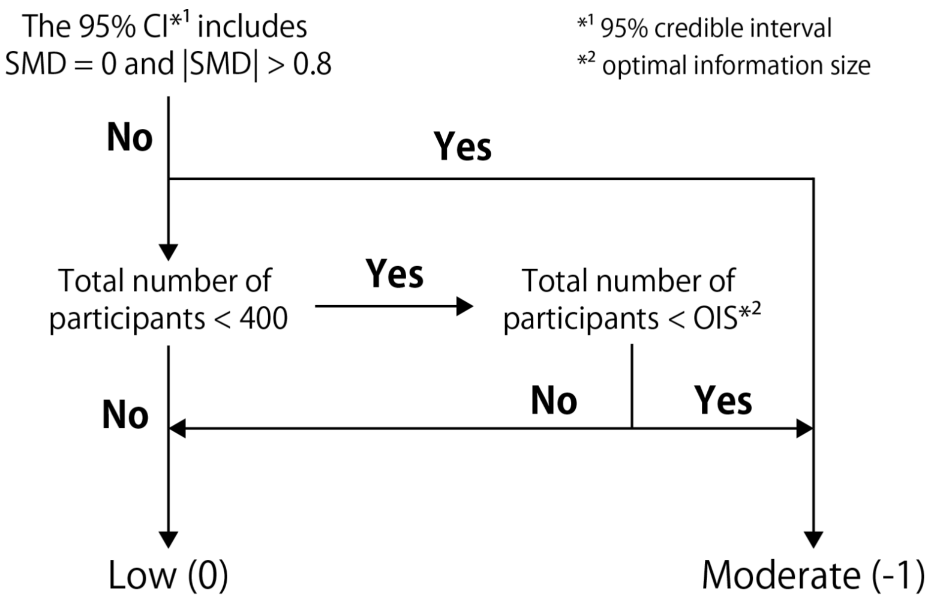


**Supplementary Figure 4**. Rating procedure for imprecision. Abbreviation: optimal information size (OIS).

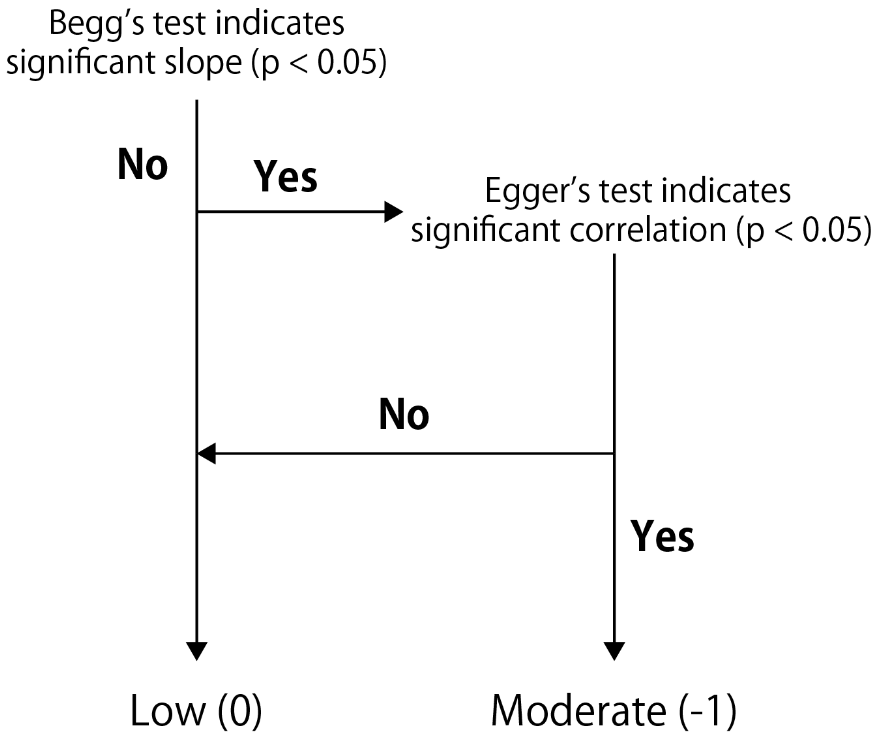


**Supplementary Figure 5.** Rating procedure for publication bias.
